# Maternal ambient air pollution exposure and risk of stillbirth in Georgia, USA

**DOI:** 10.64898/2026.01.26.26344822

**Authors:** Chen Li, Thomas W. Hsiao, Joshua L. Warren, Lyndsey A. Darrow, Matthew J. Strickland, Armistead G. Russell, Howard H. Chang

## Abstract

**Background:** Evidence suggests maternal exposure to ambient air pollution increases the risk of stillbirth, but few studies conducted in the United States have evaluated temporally varying exposures or susceptibility across gestational windows. Moreover, the generalizability of existing findings is often limited by restricted geographic coverage or reliance on selected study populations.

**Methods:** Using Georgia vital records from 2005 to 2014, we conducted a matched case-control study including 8,384 stillbirths and 33,459 live birth controls matched on maternal county of residence and conception month. We used stratified Cox proportional hazards models with time-varying covariates to estimate hazard ratios (HRs) for ten air pollutants across five exposure windows (first month, weekly, and first, second, and third trimester). Our primary analysis included all stillbirths combined, with subgroup analyses separating second and third trimester losses.

**Results:** Stillbirths had a median gestational age of 27 weeks (IQR: 6.67) compared with 38 weeks for live births (IQR: 2.13). Particulate matter showed strong associations in the second trimester exposure window for all stillbirths (PM_10_: HR = 1.07; 95% CI: 1.04, 1.11; PM_2.5_: HR = 1.05; 95% CI: 1.01, 1.09). This pattern was consistent for NO_2_ and NH_4_, which also exhibited positive associations across early and entire pregnancy exposure windows (first month, first trimester, weekly), with the strongest associations for the second trimester exposures. Associations were larger for second trimester stillbirths, whereas estimates for third trimester stillbirths were largely null or negative.

**Conclusions:** In this population-based study in Georgia, time-varying ambient air pollution exposures during pregnancy were associated with increased risk of stillbirth, particularly for second trimester exposures and for stillbirths occurring earlier in pregnancy. These findings highlight the importance of considering gestational timing when evaluating environmental risk factors for stillbirth.

**What this study adds:** This study is the first to evaluate maternal ambient air pollution exposure and stillbirth using time-varying exposures on vital records in the state of Georgia. By examining ten air pollutants across multiple gestational windows and subset analyses by timing of stillbirth, we identified second trimester susceptibility to NO_2_, PM_10_, PM_2.5_, and NH_4_. These findings highlight periods of vulnerability to ambient air pollution during pregnancy.

## Introduction

Stillbirth is defined as a pregnancy loss at or after 20 weeks of gestation.^1^ Though stillbirth is more common in low and middle-income countries, the stillbirth rate in the United States (U.S.) is higher than that of many other developed nations.^2^ In the U.S., approximately one in every 160 pregnancies results in a stillbirth, with roughly 23,000 cases occurring annually.^2^ Over the past two decades, this burden has remained relatively unchanged, corresponding to an estimated 20,000 American families a year in which a stillbirth has occurred.^3^ Experiencing a stillbirth impacts women and families in ways that reach far beyond a loss of life, including high financial cost associated with medical management,^1^ emotional and psychological stress,^3^ and health problems such as chronic pain and fatigue following the pregnancy loss.^4^

Although understanding the causes of stillbirth is critical for prevention, nearly one-third of cases remain etiologically unexplained, underscoring the need to identify modifiable risk factors to improve pregnancy outcomes, especially in the U.S.^3^ Studies have highlighted that low income, lower education attainment, and poorer health care access are associated with higher rates of stillbirth.^2^ There is an increasing body of evidence establishing the association of stillbirth with maternal ambient air pollution exposure. A recent literature review on outdoor air pollution and pregnancy loss reported that exposure to fine particulate matter (PM_2.5_), nitrogen dioxide (NO_2_), and ozone (O_3_) has consistently been associated with increased risk of stillbirth.^1^ However, only four out of the 16 studies reviewed were conducted in the U.S.^1^ Similarly, another review identified 22 studies on air pollution and stillbirth, of which only five were based in the U.S.^5^ Of the nine total studies conducted in the U.S., four of them analyzed study populations in California.

In addition to the small number of U.S. study populations in the literature, existing findings on the link between certain air pollutants and stillbirth, while relatively well-studied compared to outcomes like spontaneous abortion, still contain some unexplained discrepancies. A study conducted in New Jersey and another in California indicated PM_2.5_ exposure during any trimester, as well as across the entire pregnancy, was not significantly associated with stillbirth.^6,7^ However, a study in Ohio showed that exposure to high levels of PM_2.5_ in the third trimester of pregnancy was associated with 42% increased stillbirth risk.^8^ A United Kingdom (UK) study even observed negative associations for the first and second trimester exposure windows.^6^ In the case of O_3_, a study of pregnant women enrolled in Medicaid across the U.S. identified no association with stillbirth,^9^ yet positive associations over the entire gestational period were found in Harris County, Texas.^10^ Pollutants like nitrogen oxides (NO_x_) and PM_10_ have been investigated less frequently. The limited evidence includes one study reporting a weak negative association between NO_x_ and stillbirth,^11^ and another study reporting a positive association for PM_10_.^12^ Other pollutants had stronger agreement across studies. The association between NO_2_ and stillbirth was consistently positive,^7,13^ except for one negative association in a London study.^11^ Several studies have examined the associations between carbon monoxide (CO) and stillbirth, but none have reported any relationship.^12,14–17^

Although many studies have examined air pollution and stillbirth, important gaps and methodological limitations remain. Besides the inconsistency of the reported associations across studies and lack of data mentioned, most of the existing studies in the U.S. treat air pollution exposure as time-invariant by averaging concentrations over a specified gestational window,^6–8^ ignoring temporal variability and potential short-term fluctuations in exposure. Other studies have limited generalizability; for example, the study restricted to pregnant women enrolled in Medicaid resulted in a study population that was younger, more racially diverse, and lower-income than the general U.S. population.^9^

To add to the U.S. body of evidence on the air pollution and stillbirth association and address some of these methodological limitations, we used a time-to-event approach previously used in an analysis of air pollution and spontaneous abortion to estimate the association between maternal ambient air pollution exposure and risk of stillbirth.^18^ We conducted a matched case-control study over 2005 to 2014 with 8,384 stillbirth cases ascertained from statewide Georgia vital records. Each stillbirth case was matched to four live birth controls by maternal residential county and conception month to account for residual unmeasured spatiotemporal confounding. We used stratified Cox proportional hazards models to estimate associations between stillbirth and a comprehensive set of ten ambient particulate and gaseous pollutants (CO, NO_2_, NO_x_, O_3_, PM_10_, PM_2.5_, elemental carbon [EC], organic carbon [OC], nitrate [NO_3_], and ammonium [NH_4_]) over five different prenatal windows: the first month of pregnancy, weekly averages, and the first, second, and third trimester-specific averages.

## Method

We used vital records from the U.S. State of Georgia with an estimated pregnancy start date from January 1^st^, 2005, to February 27^th^, 2014. Georgia comprises 159 counties with substantial variation in geographic and population size. Based on the 2010 Decennial Census, the median county land area in Georgia was 898 km^2^ (range: 314 to 2,349) and median population size was 22,598 persons (range: 1,717 to 920,581 persons). Gestational age based on the clinical estimate was used to define the pregnancy start date. When unavailable, the date of the last menstrual period (LMP) was used instead (<1% of pregnancies). The conception date was then approximated as 14 days after the beginning of pregnancy. We defined stillbirth as the death of a fetus at or after 20 completed weeks of gestation. Because there were very few stillbirth events compared to live births in our dataset, we used a case-control design to improve computational efficiency while sacrificing minimal statistical efficiency. This approach has been previously applied in studies assessing the impact of air pollution on adverse pregnancy outcomes,^18,19^ where stillbirths are treated as cases and live births past 20 weeks gestational age are treated as controls. Inclusion criteria for the dataset included singleton birth status and recorded maternal residential census tract, marital status, and race/ethnicity data (Figure 1). A reported birthweight between 250g to 7500g was used as inclusion criterion only for live births.

**Figure 1.**
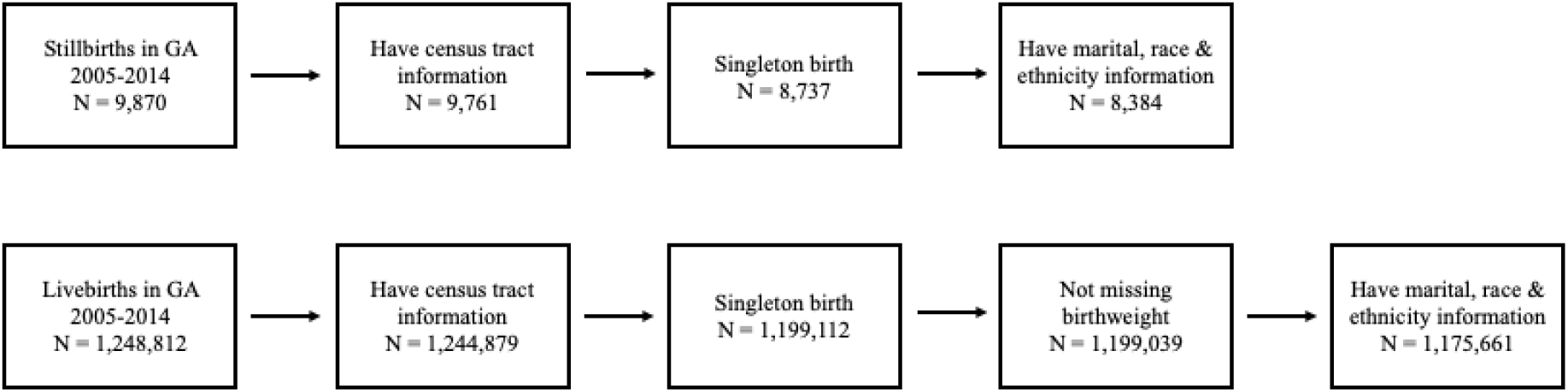
Inclusion criteria for stillbirths and live births

Air pollution exposures were estimated using a random forest model that fused outputs from the Community Multiscale Air Quality (CMAQ) chemical transport model with monitoring data from the Environmental Protection Agency’s Air Quality System (AQS). Detailed procedures for the estimates have been documented elsewhere.^20^ The estimates provided daily average pollutant concentrations on a 12 x 12 km grid, which we linked to each census tract in Georgia using spatial area-weighted averaging. In this study, we included pollutants CO, NO_x_, NO_2_, O_3_, PM_2.5_, PM_10_, and PM_2.5_ constituents NO_3_, NH_4_, EC, and OC.

For each pollutant, we assessed exposure over five different time windows: 1) the time-invariant average exposure during the first four weeks of gestation (*first month exposure*), 2) the time-varying weekly average for each gestational week from 20 weeks of gestation to the end of follow-up (*weekly exposure*), 3) the time-invariant average exposure during the first 13 weeks of gestation (*first trimester exposure*), 4) the time-varying cumulative weekly average from 20 to 27 weeks of gestation, where the cumulative exposure at the 20^th^ week was defined as the average exposure over weeks 14 to 20, with subsequent weeks updating cumulatively thereafter (*second trimester exposure*), and 5) the time-varying cumulative weekly average from 28 weeks of gestation up to the end of pregnancy (*third trimester exposure*). For the time-varying exposures, gestational age in weeks was used as the underlying time scale in the Cox model with exposures updated weekly until the end of follow-up. These exposures were selected to characterize accumulation of exposure within a time-to-event framework and therefore differ from averages computed over fixed exposure windows.

Our primary analysis analyzed all stillbirths together, but in subgroup analyses we examined stillbirths occurring in the second trimester (20-27 weeks) separately from stillbirths occurring in the third trimester (>= 28 weeks) due to potential etiologic differences between early and late stillbirths. In our matched case-control design, we adjusted for unmeasured spatiotemporal confounding (by potential factors like healthcare access, stillbirth reporting, and seasonally varying exposures) by randomly selecting four controls among all live births for each stillbirth case with matching maternal residential county and conception month. Maternal residential county was recorded at the time of delivery or stillbirth. When four matched controls could not be identified, we selected as many as available, with at least one control per case. All five exposure windows except the third trimester window were evaluated for all stillbirths and for analyses restricted to second and third trimester stillbirths. The third trimester exposure window was evaluated only among third trimester stillbirths.

To understand the extent of collinearity, we calculated Pearson’s correlation coefficient across all air pollutant-exposure window pairs. Similar to Hsiao et al.,^18^ we used a stratified Cox proportional hazards model to estimate the hazard ratio (HR) of stillbirth associated with an increase in maternal air pollution exposure across different exposure windows. In stratified Cox models, the baseline hazard function is allowed to vary across strata while assuming a common HR across all strata. We used maternal county of residence and conception month as the stratification variables to align with our matching procedure for cases and controls. Maternal county of residence was also included as a clustering variable to account for spatial correlation among observations in geographically proximate areas. For the time-varying exposure model, we have:

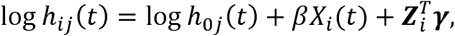

where *h*_*ij*_ (*t*) is the hazard rate on gestational week *t* for pregnancy *i* in stratum *j*, *h*_0*j*_ (*t*) is the baseline hazard rate in stratum *j*, *X*_*i*_(*t*) is the air pollution exposure that is potentially time-varying, *β* is the log HR for the pollutant of interest, ***Z***_*i*_ is the vector of confounders, and *γ* is the coefficient vector for the confounders. For the time invariant exposures like the first month and first trimester, *X*_*i*_ (*t*) reduces to *X*_*i*_.

As the risk period of stillbirth begins at 20 weeks of gestation, we considered the 20^th^ week as the starting point of observation for our survival analysis. For time-invariant exposure windows, we standardized each exposure by dividing by its corresponding IQR. For time-varying exposure windows, we calculated the IQR from the pooled weekly exposure of all individuals for each pollutant, then divided the exposure by the IQR. We adjusted for several potential confounders, including maternal age, race/ethnicity (categorized as Non-Hispanic Black, Non-Hispanic White, Hispanic or Other), marital status (married vs. unmarried), conception year (from 2005 to 2014 as a categorical variable), and tract-level median household income at the time of conception, obtained from the 2005-2009 and 2010-2014 American Community Surveys (ACS). We used penalized splines with four degrees of freedom to account for the non-linearity of HHI and maternal age.

To evaluate the impact of adjusting for temperature, we conducted sensitivity analyses in which separate Cox models were fit with weekly average minimum or maximum temperature included as covariates, specified with the same temporal resolution as the exposure. For example, sensitivity analysis models for the first month exposure window model also included the corresponding first month average daily temperature. We extracted daily temperature data from the *daymet* package in R (Thornton et al., 2022). As *daymet* does not provide the temperature data on the 366^th^ day of a leap year, we imputed it by the average temperature on the day after and the day before. We also conducted a sensitivity analysis restricting the study population to stillbirth cases and live birth controls occurring after 37 completed weeks of gestation to compare to our third trimester stillbirth results. As a further sensitivity analysis, we fitted two-pollutant models to assess the robustness of the observed single pollutant associations in the main analysis. We focused on pollutant pairs commonly considered in co-pollutant air pollution analyses including NO_2_ adjusted for PM_2.5_, and O_3_ and PM_2.5_ adjusted for each other. Finally, to investigate any differences in associations by gestational age at stillbirth, we ran a secondary analysis comparing associations among preterm stillbirths (<37 completed weeks gestation) with those in term stillbirths (37-42 completed weeks). All other specifications were unchanged from the main analysis.

## Results

Table 1 shows the demographic characteristics of the study population. There were 8,384 stillbirths, with an average maternal age of 27.3 years (SD: 6.65 years) and average gestational age of 27.2 weeks (SD: 6.67 weeks). There were 33,459 live births included, with an average maternal age of 27.2 years (SD: 6.65 years) and average gestational age of 38.4 weeks (SD: 2.13 weeks). Compared with the controls, individuals who experienced stillbirth tended to reside in census tracts with lower median household income (46,700 vs. 50,000 USD) and a higher proportion of Black residents (54.8% vs. 37.1%). The proportion married was also lower among stillbirths (41.0% vs. 53.9%). Based on the distribution of gestational age at stillbirth, most stillbirths occurred between the 20th and 24th weeks of gestation, with a smaller peak between the 35th and 39th weeks (Figure 2). For most of the air pollutants, the stillbirth group had slightly higher exposure levels than the control group (Table 2). Some of the air pollutants were strongly positively correlated, such as the pairs of NO_3_ and CO, NO_3_ and NO_x_, and PM_10_ and PM_2.5_ (Table S1). Within each pollutant, exposure levels in the first and second trimesters were highly correlated, whereas correlations between the first and third trimesters, as well as between the second and third trimesters, were relatively weak (Table S2).

**Figure 2.**
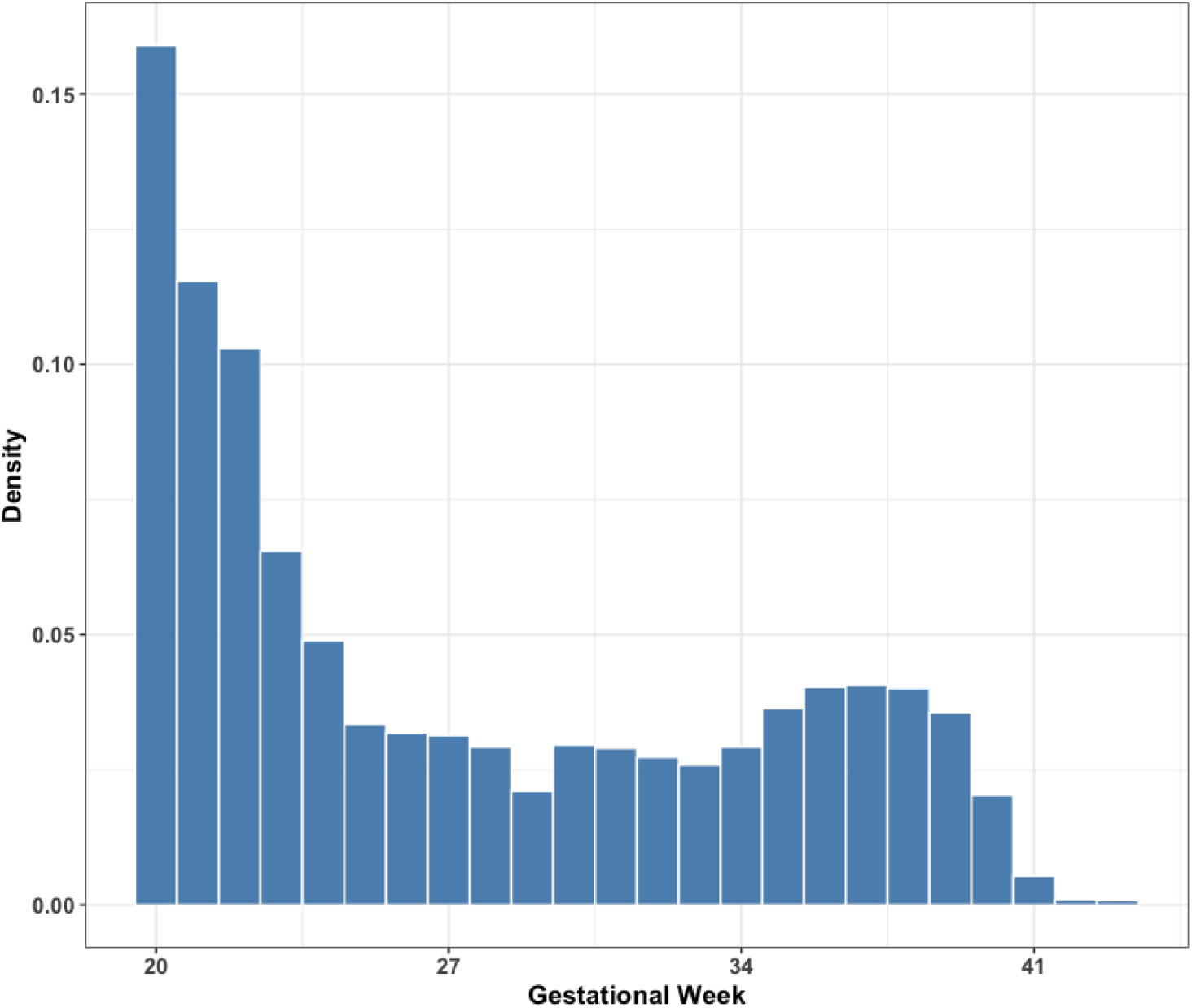
The gestational age (by week) distribution for all individuals with stillbirths from 2005 to 2014.

**Table 1.** Baseline characteristics of stillbirth cases and live birth controls from the case-control analysi for the first month, weekly, first trimester, second trimester and the third trimester exposure, Georgia, United States, 2005-2014.

| Characteristics | Live birth | Stillbirth | Total |
| --- | --- | --- | --- |
| n | 33459 | 8384 | 41843 |
| Gestational age (weeks) | 38.41 (2.13) | 27.24 (6.67) | 36.17 (5.71) |
| Maternal age (years) | 27.18 (6.07) | 27.30 (6.65) | 27.21 (6.19) |
| Maternal race (%) |  |  |  |
| White | 14053 (42.0) | 2665 (31.8) | 16718 (40.0) |
| Black | 12404 (37.1) | 4598 (54.8) | 17002 (40.6) |
| Hispanic | 4934 (14.7) | 850 (10.1) | 5784 (13.8) |
| Other | 2068 (6.2) | 271 (3.2) | 2339 (5.6) |
| Mother married (%) | 18029 (53.9) | 3441 (41.0) | 21470 (51.3) |
| Tract-level median household income (\$) | 50044.24 (22078.37) | 46736.46 (20092.32) | 49381.46 (21735.14) |
| Tract-level percent below poverty (%) | 0.19 (0.12) | 0.20 (0.12) | 0.19 (0.12) |
| Conception year (%) |  |  |  |
| 2004 - 2009 | 18776 (56.1) | 4702 (56.1) | 23478 (56.1) |
| 2010 - 2014 | 14683 (43.9) | 3682 (43.9) | 18365 (43.9) |
| Conception season<sup>a</sup> (%) |  |  |  |
| Spring | 8343 (24.9) | 2089 (24.9) | 10432 (24.9) |
| Summer | 7600 (22.7) | 1906 (22.7) | 9506 (22.7) |
| Fall | 8479 (25.3) | 2124 (25.3) | 10603 (25.3) |
| Winter | 9037 (27.0) | 2265 (27.0) | 11302 (27.0) |

<sup>a</sup>Seasons are defined as Spring (March-May), Summer (June-Aug.t), Fall (Sept.-Nov.), and Winter (Dec.-Feb.)

**Table 2.** Mean (SD) of individual-level weekly average pollutants exposure for live birth controls, stillbirth cases, and total combined.

| Air pollutants | Non-stillbirth | Stillbirth | Total |
| --- | --- | --- | --- |
| CO (ppm) | 0.59 (0.27) | 0.60 (0.28) | 0.59 (0.27) |
| NO2 (ug/m<sup>3</sup>) | 16.36 (8.32) | 16.50 (8.47) | 16.39 (8.35) |
| NOx (ppb) | 30.47 (20.08) | 30.78 (21.10) | 30.53 (20.29) |
| O3 (ppb) | 41.11 (4.46) | 41.11 (3.94) | 41.11 (6.09) |
| PM10 (ug/m<sup>3</sup>) | 18.46 (3.13) | 18.53 (3.66) | 18.48 (3.24) |
| PM2.5 (ug/m<sup>3</sup>) | 11.36 (2.43) | 11.42 (2.63) | 11.37 (2.47) |
| EC (ug/m<sup>3</sup>) | 0.62 (0.25) | 0.62 (0.26) | 0.62 (0.25) |
| OC (ug/m<sup>3</sup>) | 3.24 (1.49) | 3.27 (1.52) | 3.24 (1.49) |
| NO3 (ug/m<sup>3</sup>) | 0.86 (0.33) | 0.86 (0.39) | 0.86 (0.35) |
| NH4 (ug/m<sup>3</sup>) | 0.96 (0.46) | 0.97 (0.47) | 0.96 (0.46) |

Figure 3 presents the estimated HRs and 95% confidence intervals for stillbirth per IQR increase for each air pollutant during the weekly, first month, first trimester, and second trimester exposure windows. The IQRs used for each pollutant-exposure window are reported in Table 3. Results for the third trimester exposure window can be found in Figure 4, and all numerical values are reported in Table S3. Among the associations across different exposure windows for each air pollutant, most of the significant positive associations were observed for the second trimester exposure window. For the second trimester exposure window, we observed consistent positive associations between all stillbirth cases and NO_2_ (HR = 1.064, 95% CI: 1.002- 1.131), PM_10_ (HR = 1.073, CI: 1.041 - 1.106), PM_2.5_ (HR = 1.045, CI: 1.006 - 1.085), NH_4_ (HR = 1.058, CI: 1.022 - 1.095) and CO (HR = 1.079, CI: 0.976 - 1.193). There were relatively fewer positive associations observed for the first month and first trimester exposures, regardless of stillbirth subgroup. However, apart from the first month exposure, the associations between time to stillbirth in the third trimester and O_3_ exposure windows were all positive, while those of most of the other air pollutants were null or negative. The estimate for the third trimester O_3_ exposure was particularly high (HR = 1.12, CI: 1.01, 1.24) (Table S3).

**Figure 3.**
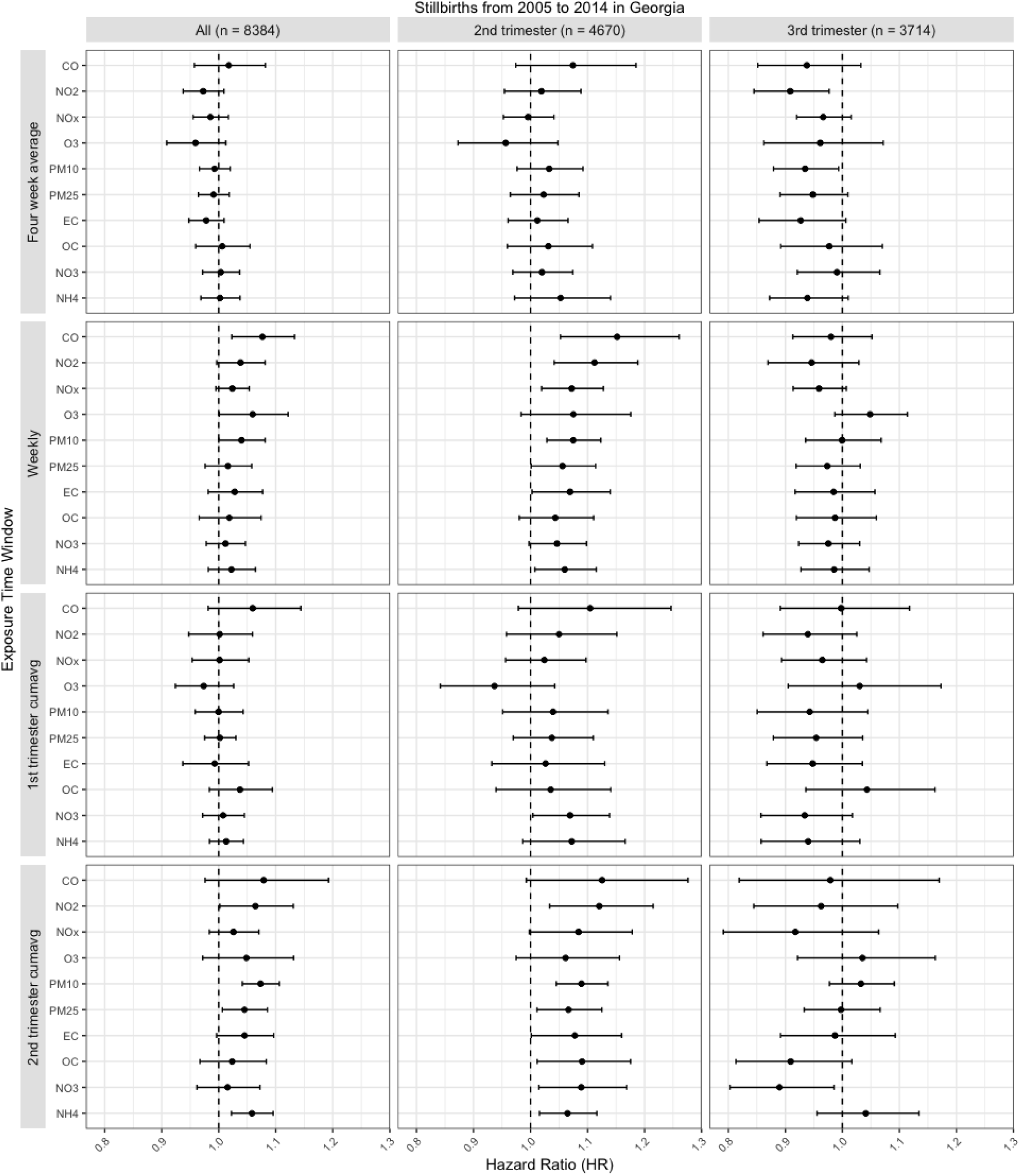
Hazard ratio (HR) estimates per unit IQR increase and 95% CIs for all ten pollutants during four exposure windows (the first month, weekly, first trimester, and second trimester exposure) for all stillbirths, second trimester stillbirths, and third trimester stillbirths. Estimates are adjusted for individual-level maternal age, marital status, race & ethnicity, conception year, and tract-level median household income.

**Figure 4.**
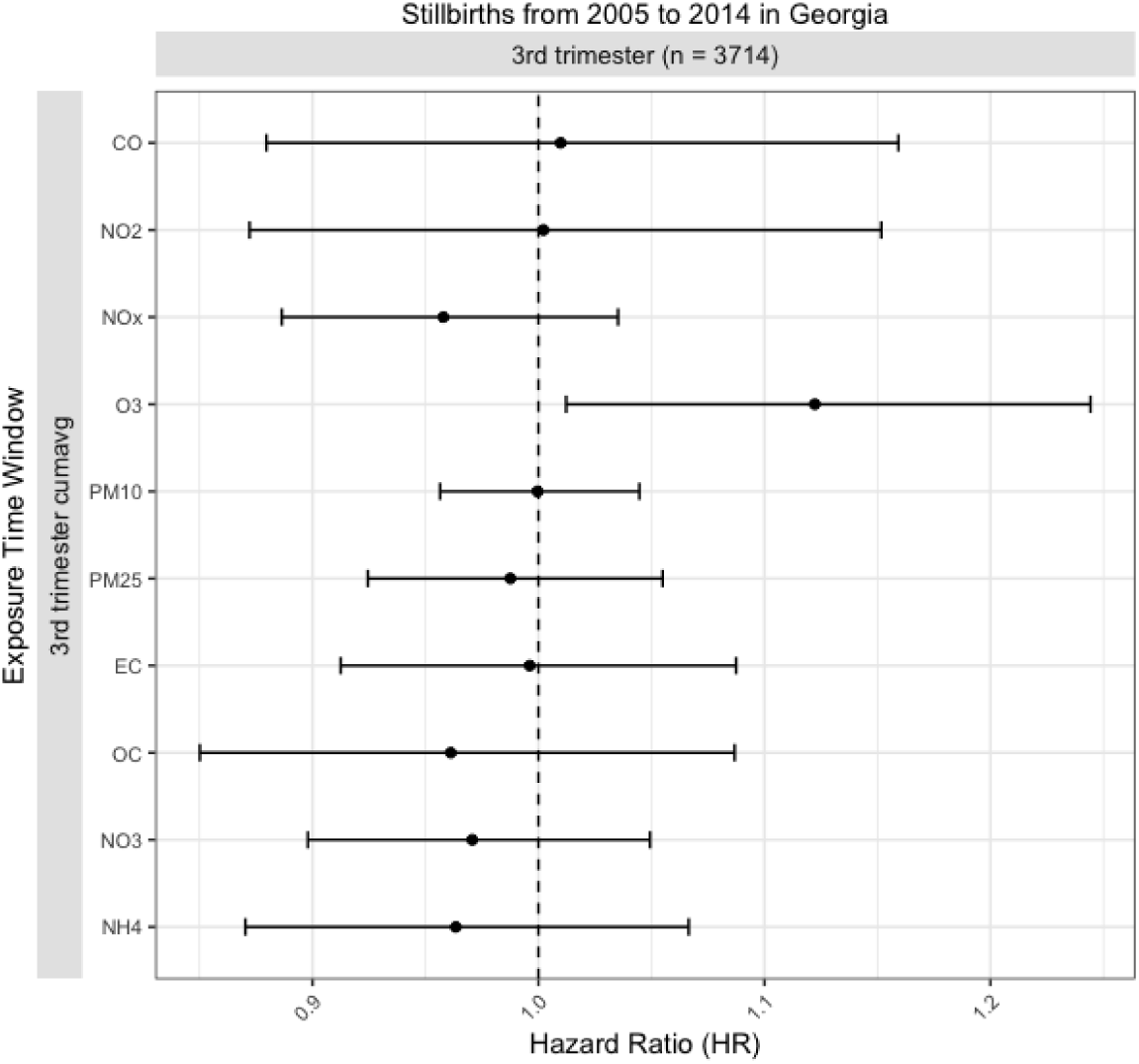
Hazard ratio (HR) estimates per unit IQR increase and 95% CI’s for all ten pollutants during the third trimester exposure window for stillbirths that occurred in the third trimester. Estimates are adjusted for individual-level maternal age, marital status, race & ethnicity, conception year, and tract-level median household income.

**Table 3.** IQR for each pollutant-exposure window used to report HRs.

| Air pollutants | Four week average | Weekly | 1<sup>st</sup> trimester cumavg | 2<sup>nd</sup> trimester cumavg | 3<sup>rd</sup> trimester cumavg |
| --- | --- | --- | --- | --- | --- |
| CO (ppm) | 0.43 | 0.41 | 0.43 | 0.42 | 0.4 |
| NO2 (ug/m<sup>3</sup>) | 13.84 | 13.71 | 13.79 | 13.63 | 13.57 |
| NOx (ppb) | 31.31 | 29.57 | 31.72 | 30.47 | 30.48 |
| O3 (ppb) | 14.89 | 15.36 | 13.4 | 13.17 | 13.85 |
| PM10 (ug/m<sup>3</sup>) | 6.36 | 6.9 | 5.67 | 5.86 | 5.92 |
| PM2.5 (ug/m<sup>3</sup>) | 4.05 | 4.54 | 3.71 | 3.82 | 3.85 |
| EC (ug/m<sup>3</sup>) | 0.42 | 0.44 | 0.42 | 0.42 | 0.43 |
| OC (ug/m<sup>3</sup>) | 2.64 | 2.56 | 2.71 | 2.66 | 2.59 |
| NO3 (ug/m<sup>3</sup>) | 0.76 | 0.75 | 0.7 | 0.7 | 0.72 |
| NH4 (ug/m<sup>3</sup>) | 0.75 | 0.74 | 0.74 | 0.74 | 0.73 |

Comparing the results from different stillbirth population groups, the associations for all stillbirths were close to null or slightly positive, while the HRs of the second trimester stillbirths were strongly positive, but null or negative for the third trimester stillbirths. For example, the HRs for all stillbirths during the second trimester exposure window were all positive but less than 1.10, whereas the HRs of CO and NO_2_ were above 1.10 and the HRs for all other air pollutants exhibited increases of varying magnitudes when the second trimester only stillbirths were analyzed. However, the HRs decreased to null, even to negative in the third trimester only analysis. To investigate whether this finding could be due to the shorter duration of third trimester stillbirths, we examined a sensitivity analysis restricted to full-term stillbirths and live births (> 37 weeks). We found that all HRs remained near null except for a positive HR for NO_2_ during the second trimester window (Figure S1, Table S4). Additionally, similar results were observed for most of the pollutants during the weekly and first trimester exposure windows. CO always had the largest magnitude of HRs across all the air pollutants. Specifically, we observed positive association between all stillbirth cases and CO during the first month (HR = 1.018, CI: 0.957 – 1.082), weekly (HR = 1.077, CI: 1.023, 1.133), first trimester (HR = 1.060, CI: 0.981, 1.144), and second trimester (HR = 1.079, CI: 0.976, 1.193). The HRs of CO all increased when only analyzing the second trimester only stillbirth cases, to 1.074 (CI: 0.974, 1.185) for first month, 1.152 (CI: 1.053, 1.261) for weekly, 1.105 (CI: 0.979, 1.247) for first trimester, and 1.126 (CI: 0.993, 1.276) for second trimester. However, the associations between third trimester only stillbirths and CO were all negative or close to null.

Comparing the first and second trimester exposure windows, the associations for all three stillbirth population groups share similar patterns: many of them had the same direction of HRs within each pollutant, and the relative ordering of pollutant-specific HRs was consistent across different exposure windows. For example, we observed the same HRs in decreasing order to be NO_2_, NO_x_, PM_10_, then O_3_ in the second trimester cases analysis. In our sensitivity analysis, we found that adjustment for weekly average maximum (Figure S2, Table S5) or minimum temperature (Figure S3, Table S6) had minimal impact on the estimated HRs. The sensitivity analysis for the two pollutant models produced results that were broadly similar to those from the corresponding single pollutant models (Figure S4, Table S7). Positive associations for NO_2_ were slightly attenuated after adjustment for PM_2.5_, whereas results for O_3_ and PM_2.5_ showed greater variability across exposure windows and stillbirth subgroups with no clear pattern. In the analysis stratified by gestational age at stillbirth, associations were generally stronger among preterm than term stillbirths (Figure S5, Table S8). In contrast, estimates among term stillbirths were often closer to the null, although some pollutants showed elevated HRs for later second and third trimester exposures. These estimates were based on a smaller number of events and had wider 95% confidence intervals than the corresponding preterm stillbirth estimates.

## Discussion

We conducted a matched case-control study analyzing 8,384 stillbirth cases from 2005 to 2014 in the state of Georgia to examine the association between a comprehensive set of ten air pollutants and stillbirth. Cox proportional hazard models with time-varying exposure were used to account for the weekly fluctuations of air pollutants. We identified positive HRs for stillbirth per IQR increase of NO_2_, PM_10_, PM_2.5_, and NH_4_ during the second trimester exposure window for all stillbirth cases. The magnitude of HRs was higher for stillbirth cases occurring in the earlier second trimester and preterm periods, compared to those of all stillbirths, third trimester, and full term stillbirths. Estimated HRs for third trimester and full term stillbirths were mostly null or even negative; these findings warrant careful interpretation, which we discuss later in this section.

We observed positive associations between PM_2.5_ and stillbirth during the second trimester exposure window and the entire pregnancy for all stillbirth cases, which aligns with prior literature. Positive associations between stillbirth and both average annual PM_2.5_ concentration and average monthly PM_2.5_ concentration were observed in the United States.^21,22^ A recent study conducted in Guangzhou, China identified positive HRs for stillbirths with each 10 μg/m3 increase in PM_2.5_ concentration during the second trimester and the entire pregnancy.^23^ However, this study also found positive associations during the first trimester and the third trimester, whereas we observed null associations in these periods. Similarly, two earlier studies in China found positive associations for exposure in all three trimesters in Wuhan^24^ and the Coastal area of China.^25^ In the most recent Guangzhou study, which also had the largest cohort (1,273,924 participants and 3,150 stillbirths), the strongest associations were observed during the second trimester, followed by the first and then the third trimester. This ordering was similar to what we observed in our analysis and was also supported by an Australian study identifying the critical windows of susceptibility to PM_2.5_ for birth loss.^26^ The study found that the 5^th^ to 6^th^ gestational months were the most sensitive, corresponding to the second trimester exposure window in our study. Our findings, together with those from the Australian and Guangzhou studies, indicate that pregnant women may be more susceptible to PM_2.5_ exposure during the second trimester, providing insight into the timing of potential interventions and underlying mechanisms. Pollutants PM_10_, NH_4_, and NO_2_ exhibited the same pattern as PM_2.5_, suggesting increased susceptibility to these pollutants during the same period.

While our PM_2.5_ findings were generally consistent with the studies listed above, notable discrepancies remain. In the three studies from China mentioned above,^23–25^ they found positive associations for the first and third trimester exposure windows but our results were null for these two periods. Unlike our analysis, these three studies defined trimester-specific exposure as the average of all daily pollutant concentrations. Additionally, the average exposure to PM_2.5_ for these three studies were all over 30 μg/m^3^, much higher than levels in Georgia from 2005 to 2014.^27^ The time invariance of their air pollution exposure and the elevated baseline exposure level may be related to the difference in estimates. Nevertheless, a California cohort study found null associations for all three trimesters and the entire pregnancy,^28^ and used the mean concentration in each trimester and the whole period as well. Unlike our stratified Cox proportional hazards model, the logistic regression models used in that study did not account for residual spatial confounding (e.g., by factors like access to healthcare, stillbirth reporting, and seasonally varying exposures). However, they did control for extra individual-level confounders including maternal education level and the sex of the fetus but did not control for marital status and tract-level median household income.

For NO_2_, we observed positive association during the second exposure window, a finding not previously reported in the literature. Studies conducted in London (UK), Jiangsu (China), Wuhan (China), California, and New Jersey all reported null associations during this window.^29–32^ Other studies identified associations in other windows: the New Jersey study observed positive associations during the first trimester and entire pregnancy,^31^ the study in California also observed positive associations during the entire pregnancy,^33^ and the study in Wuhan identified positive associations during the third trimester.^29^ In contrast, our findings for these windows were null. These inconsistencies may reflect geographical heterogeneity in the air pollutant exposure levels. For example, the average daily NO_2_ concentration for any area in Georgia in our study was less than 35 ppb,^27^ while the mean NO_2_ concentrations in the California study were 36.24 ppb and 35.94 for stillbirths and livebirths,^33^ all greater than 35 ppb. Methodological differences may also be a factor. Prior studies relied on simple logistic regression with time-invariant exposures, whereas we used a Cox model with time-varying exposure to better account for temporal variability in pollutant exposure during pregnancy.

For CO, we observed consistently elevated HRs during the first, second, and third trimester and the entire pregnancy exposure window (weekly exposure), although statistical significance was only reached during the entire pregnancy. Our findings were similar to a study in New Jersey reporting significant positive associations only during the third trimester exposure window and the entire pregnancy.^34^ In terms of toxicological mechanisms, CO’s effects are more direct and potent than those of other air pollutants: binding hemoglobin for both mothers and fetuses to prevent the circulation of O_2_, which could cause severe damage, including intrauterine hypoxia, neurological damage, and even fetal death.^35,36^ While this may partly explain the elevated HRs for CO compared to other pollutants, the concentrations necessary to reach lethal levels are typically much higher than those observed in ambient air pollution. In our analysis, the confidence intervals around our CO estimates were wider relative to those of the other pollutants. This observation suggests higher uncertainty in the estimated CO exposure-response relationship, which is further supported by the similar CO exposure distributions observed between the stillbirth and non-stillbirth populations. This finding may be consistent with unexplained heterogeneity in the study population. Such variability in outcomes within the same exposure level may be explained by differential susceptibility to CO across individuals. For example, persons with COPD, anemia, cerebrovascular disease (CBD), heart failure, multiple co-morbidities, and persons of older age (≥ 60 years) are at increased risk of adverse effects by exposure to CO.^37^ An alternative explanation is that CO may act as a proxy for other components of mobile-source pollution that were not directly measured or are more spatially heterogeneous, which could contribute to both the elevated point estimates and increased uncertainty.

A surprising finding from our study was a high estimated HR for O_3_ during the third trimester exposure window, while other pollutants were observed as null or negative. Two studies, in Yancheng, China^39^ and the state of California^38^ also reported positive associations between O_3_ and stillbirth during the third trimester exposure window but null or negative associations for other pollutants including NO_2_, CO, SO_2_, and PM_10_. However, two other studies from Wuhan^40^ and Taiwan^41^ reported null associations between O_3_ and stillbirth for all three trimesters and the entire pregnancy. One study in London identified positive associations during the first and second trimesters but null for the third trimester.^42^ Earlier studies investigating the association between O_3_ and stillbirth only focused on exposure during the entire pregnancy.^43,44^ Though those studies reported consistent positive associations in alignment with our findings, they did not explore the variability of susceptibility across different trimesters. Beyond the stillbirth literature, a recent nationwide U.S. study linked monthly ozone exposure to increased postneonatal infant mortality due to sudden infant death syndrome (SIDS).^45^ While SIDS and stillbirth are distinct mortality outcomes, these findings provide additional evidence that ozone exposure may have adverse effects on early-life survival. More studies are needed to understand the association between O_3_ and stillbirth during different gestational windows.

Additionally, we identified a pattern shared by most pollutants. HRs for earlier periods like second trimester and preterm stillbirth cases were more positive than those for all stillbirth cases, while the HRs for the third trimester and full term stillbirth cases were null or negative. Similar patterns of stronger associations earlier in pregnancy and attenuated later in pregnancy have been reported previously. We reported similar findings in a previous study^27^ estimating second trimester HRs for the air pollution-spontaneous abortion link. According to a recent systematic review,^46^ ten of twelve studies evaluating exposure across the whole pregnancy and four of five examining the first trimester reported positive associations with stillbirth, while only two of five studies focusing on the third trimester reported positive associations. One explanation could be that pregnant women are more likely to be cautious of environmental exposures (e.g., reduce frequency and duration of outdoor activities^47^) and often take other perinatal care precautions more seriously during the late stages of pregnancy, leading to lower risk during these periods.^26^ Another possible explanation is bias due to a “depletion-of-susceptibles.” This bias arises when relatively high-risk individuals, those susceptible to stillbirth, exit the risk set during the second trimester.^48^ As a result, third trimester stillbirths consist of more resistant individuals and observed decreases in later term HRs may be due to a change in the at-risk population rather than effect size. This phenomenon may also occur earlier at the 20-week threshold, when stillbirth first becomes observable, because pregnancies most vulnerable to earlier exposure may have ended before stillbirth is defined and would instead be classified as miscarriages. The underlying cause of this pattern remains uncertain. Further research and methodology are needed to clarify whether the difference in effect estimates between earlier and later pregnancy loss is due to selection bias or a true biological mechanism.

Our study has several strengths. First, we are, to the best of our knowledge, the first study to investigate the relationship between stillbirth and air pollution not only across different exposure windows, but also by different subgroups of stillbirth cases which may have different etiologies. This allows for a detailed analysis of the relationship between air pollution and stillbirth throughout various stages of pregnancy, meanwhile providing new insights into potential temporal heterogeneity in vulnerability and underlying etiological mechanisms across gestational periods. Second, unlike previous studies that used average air pollutant exposure over a fixed time window, we utilized a stratified Cox model with time-varying exposure to capture temporal fluctuations in air pollution. Our model also accounted for variation from maternal residential county by the clustering variable and potential unmeasured confounding by the matching procedure and stratification. Third, we add to the U.S. air pollution-stillbirth literature by using vital records from the state of Georgia. Since stillbirths are by law a reportable event in Georgia, our data provide comprehensive statewide coverage. This dataset enabled the assessment of air pollution-stillbirth associations with broad geographic coverage and a large case count. Finally, our study covered a wide range of air pollutants. We included commonly studied air pollutants from previous research, such as PM_2.5_, PM_10_, O_3_, CO, and NO_2_, to build upon existing evidence. In addition, we examined less frequently analyzed pollutants, including NOD, EC, NOD, NHD, and OC, thereby laying the groundwork for future investigations.

Our study also has limitations. First, our exposure assessment may contain measurement error at the individual level and our model does not account for this uncertainty.^50^ We assigned exposures by census tract with a data product available at a 12km x 12km geographic resolution, potentially ignoring both personal and within-tract variation. Moreover, the exposures only measured outdoor concentrations, and we did not have data on maternal mobility available to account for exposure occurring outside the residential address recorded at delivery. Nevertheless, previous studies have suggested that risk estimates for ambient air pollution are often robust to exposure measurement error.^51–53^ In addition, while personal measurements better reflect an individual’s true exposure, they can also be influenced by individual behaviors that may introduce additional confounding or reverse causation. Although our area-level exposures provide less precise measurement, they may be less sensitive to these individual-level factors.^54^ Second, several key maternal health behavior adjustment variables, including tobacco and alcohol use, were missing at high rates and therefore unavailable. The absence of behavioral data may introduce bias in our estimates. However, to mitigate this bias, we were able to adjust for tract-level household income, which has been found to be related to a large number of poor health behaviors linked to adverse pregnancy outcomes.^55^ Third, although the evaluation of ten pollutants across five exposure windows provides a comprehensive assessment of potential associations, it also results in a large number of statistical comparisons, warranting cautious interpretation of the estimated HRs. Lastly, unmeasured residual confounding remains a possibility, which we sought to address through our spatiotemporal matching procedure.

## Data and code

Vital records data can be requested from the Georgia Department of Health through the Public Health Information Portal. The monitoring and CMAQ air quality data are publicly available from the US Environmental Protection Agency. All code for this analysis is available at https://github.com/chenli488/Air-Pollution-and-Stillbirth.

## Acknowledgments

None.

