## Supplementary Material for "Maternal ambient air pollution exposure and risk of stillbirth in Georgia, USA"

**Table S1**. Pearson Correlation Coefficients matrix between weekly levels of all ten pollutants from 2005 to 2014.

|  | CO | NO3 | NOx | O3 | PM10 | PM2.5 | EC | OC | NO3 | NH3 |
| --- | --- | --- | --- | --- | --- | --- | --- | --- | --- | --- |
| CO | 1 | 0.812 | 0.786 | -0.028 | 0.397 | 0.509 | 0.744 | 0.551 | 0.433 | 0.481 |
| NO2 | 0.812 | 1 | 0.888 | -0.01 | 0.314 | 0.407 | 0.762 | 0.358 | 0.47 | 0.358 |
| NOx | 0.786 | 0.888 | 1 | -0.211 | 0.184 | 0.273 | 0.752 | 0.29 | 0.48 | 0.211 |
| O3 | -0.028 | -0.01 | -0.211 | 1 | 0.589 | 0.56 | 0.082 | 0.245 | -0.352 | 0.393 |
| PM10 | 0.397 | 0.314 | 0.184 | 0.589 | 1 | 0.817 | 0.444 | 0.527 | -0.039 | 0.645 |
| PM2.5 | 0.509 | 0.407 | 0.273 | 0.56 | 0.817 | 1 | 0.542 | 0.671 | 0.148 | 0.798 |
| EC | 0.744 | 0.762 | 0.752 | 0.082 | 0.444 | 0.542 | 1 | 0.531 | 0.363 | 0.384 |
| OC | 0.551 | 0.358 | 0.29 | 0.245 | 0.527 | 0.671 | 0.531 | 1 | 0.3 | 0.604 |
| NO3 | 0.433 | 0.47 | 0.48 | -0.352 | -0.039 | 0.148 | 0.363 | 0.3 | 1 | 0.315 |
| NH4 | 0.481 | 0.358 | 0.211 | 0.393 | 0.645 | 0.798 | 0.384 | 0.604 | 0.315 | 1 |

**Table S2**. Pearson Correlation Coefficients between different exposure windows within each of ten air pollutants.

| Pollutants | Cor (T1, T2) | Cor (T1, T3) | Cor (T2, T3) |
| --- | --- | --- | --- |
| CO | 0.9414 | -0.0008 | -0.0019 |
| NO2 | 0.9166 | -0.0007 | -0.0031 |
| NOx | 0.8034 | 0.0006 | 0.0005 |
| O3 | 0.0647 | -0.0034 | -0.0028 |
| PM10 | 0.4445 | 0.0022 | -0.0012 |
| PM2.5 | 0.631 | 0.0022 | 0.0033 |
| EC | 0.8264 | 0.0047 | 0.0064 |
| OC | 0.8841 | 0.0034 | 0.0042 |
| NO3 | 0.3709 | -0.0016 | -0.0081 |
| NH4 | 0.8104 | 0.0022 | 0.0045 |

We also constructed the same table for stillbirths with gestational age >=30 weeks. Because many stillbirths have a short third trimester, correlations between third trimester exposure and earlier trimesters can seem low. Restricting to later stillbirths reduces can give a clearer depiction of trimester correlations.

| Pollutants | Cor (T1, T2) | Cor (T1, T3) | Cor (T2, T3) |
| --- | --- | --- | --- |
| CO | 0.9407 | 0.8974 | 0.9421 |
| NO2 | 0.9157 | 0.8572 | 0.9247 |
| NOx | 0.7995 | 0.6422 | 0.8128 |
| O3 | 0.0337 | -0.6803 | 0.1081 |
| PM10 | 0.431 | -0.0064 | 0.4763 |
| PM2.5 | 0.6257 | 0.3857 | 0.6451 |
| EC | 0.8266 | 0.7208 | 0.8375 |
| OC | 0.8857 | 0.8307 | 0.8868 |
| NO3 | 0.3544 | -0.1123 | 0.4154 |
| NH4 | 0.8082 | 0.6802 | 0.8206 |

**Table S3**. Hazard ratio (HR) estimates per unit IQR increase and 95% CI's for all ten pollutants during five exposure windows (the first month, weekly exposure, the first trimester exposure, the second trimester exposure, and the third trimester exposure) for all stillbirths, stillbirths that happened in the second trimester, and stillbirths that happened in the third trimester. Estimates are adjusted for individual-level maternal age, marital status, race & ethnicity, conception year, and tract-level median household income.

| Exposure time window | Pollutants | All stillbirth (n=8384) | 2nd trimester (n=4670) | 3rd trimester (n=3714) |
| --- | --- | --- | --- | --- |
| The first month | CO | 1.018 (0.957, 1.082) | 1.074 (0.974, 1.185) | 0.938 (0.852, 1.032) |
|  | NO2 | 0.973 (0.938, 1.009) | 1.019 (0.954, 1.088) | 0.908 (0.845, 0.977) |
|  | NOx | 0.985 (0.955, 1.017) | 0.996 (0.952, 1.041) | 0.966 (0.920, 1.015) |
|  | O3 | 0.959 (0.909, 1.012) | 0.956 (0.873, 1.048) | 0.961 (0.862, 1.072) |
|  | PM10 | 0.993 (0.966, 1.020) | 1.033 (0.976, 1.092) | 0.935 (0.879, 0.993) |
|  | PM2.5 | 0.991 (0.964, 1.018) | 1.023 (0.965, 1.085) | 0.948 (0.890, 1.010) |
|  | EC | 0.978 (0.948, 1.009) | 1.012 (0.961, 1.066) | 0.927 (0.854, 1.006) |
|  | OC | 1.006 (0.960, 1.055) | 1.031 (0.959, 1.109) | 0.977 (0.892, 1.070) |
|  | NO3 | 1.004 (0.972, 1.037) | 1.020 (0.969, 1.074) | 0.991 (0.921, 1.066) |
|  | NH4 | 1.003 (0.969, 1.037) | 1.053 (0.972, 1.140) | 0.939 (0.872, 1.010) |
| Weekly | CO | 1.077 (1.023, 1.133) | 1.152 (1.053, 1.261) | 0.980 (0.913, 1.052) |
|  | NO2 | 1.038 (0.997, 1.081) | 1.112 (1.041, 1.188) | 0.946 (0.870, 1.029) |
|  | NOx | 1.024 (0.995, 1.053) | 1.072 (1.019, 1.128) | 0.959 (0.914, 1.007) |
|  | O3 | 1.059 (1.001, 1.122) | 1.075 (0.983, 1.176) | 1.049 (0.987, 1.114) |
|  | PM10 | 1.040 (1.000, 1.081) | 1.075 (1.029, 1.123) | 0.999 (0.936, 1.068) |
|  | PM2.5 | 1.016 (0.976, 1.058) | 1.056 (1.001, 1.114) | 0.973 (0.919, 1.031) |
|  | EC | 1.028 (0.982, 1.077) | 1.069 (1.003, 1.140) | 0.985 (0.917, 1.057) |
|  | OC | 1.019 (0.966, 1.074) | 1.043 (0.980, 1.111) | 0.987 (0.919, 1.060) |
|  | NO3 | 1.012 (0.978, 1.047) | 1.046 (0.997, 1.098) | 0.975 (0.923, 1.030) |
|  | NH4 | 1.022 (0.982, 1.064) | 1.060 (1.007, 1.115) | 0.985 (0.927, 1.047) |
| The first trimester cumulative average | CO | 1.060 (0.981, 1.144) | 1.105 (0.979, 1.247) | 0.998 (0.891, 1.118) |
|  | NO2 | 1.002 (0.947, 1.059) | 1.050 (0.958, 1.151) | 0.939 (0.861, 1.025) |
|  | NOx | 1.002 (0.953, 1.053) | 1.024 (0.956, 1.097) | 0.965 (0.893, 1.042) |
|  | O3 | 0.974 (0.924, 1.026) | 0.937 (0.842, 1.042) | 1.030 (0.905, 1.173) |
|  | PM10 | 1.000 (0.959, 1.043) | 1.039 (0.951, 1.136) | 0.943 (0.851, 1.045) |
|  | PM2.5 | 1.002 (0.975, 1.030) | 1.037 (0.970, 1.110) | 0.954 (0.879, 1.036) |
|  | EC | 0.993 (0.937, 1.052) | 1.026 (0.932, 1.130) | 0.948 (0.868, 1.035) |
|  | OC | 1.037 (0.984, 1.094) | 1.035 (0.940, 1.141) | 1.043 (0.936, 1.162) |
|  | NO3 | 1.008 (0.972, 1.045) | 1.069 (1.004, 1.139) | 0.934 (0.857, 1.018) |
|  | NH4 | 1.013 (0.984, 1.043) | 1.072 (0.986, 1.166) | 0.940 (0.857, 1.031) |
| The second trimester cumulative average | CO | 1.079 (0.976, 1.193) | 1.126 (0.993, 1.276) | 0.979 (0.819, 1.170) |
|  | NO2 | 1.064 (1.002, 1.131) | 1.121 (1.033, 1.215) | 0.963 (0.845, 1.097) |
|  | NOx | 1.026 (0.983, 1.070) | 1.084 (0.998, 1.178) | 0.917 (0.791, 1.064) |
|  | O3 | 1.049 (0.972, 1.131) | 1.062 (0.975, 1.156) | 1.035 (0.921, 1.163) |
|  | PM10 | 1.073 (1.041, 1.106) | 1.089 (1.045, 1.136) | 1.033 (0.977, 1.091) |
|  | PM2.5 | 1.045 (1.006, 1.085) | 1.067 (1.011, 1.125) | 0.997 (0.933, 1.066) |
|  | EC | 1.045 (0.997, 1.096) | 1.078 (1.001, 1.160) | 0.987 (0.891, 1.093) |
|  | OC | 1.024 (0.967, 1.083) | 1.090 (1.011, 1.175) | 0.909 (0.813, 1.017) |
|  | NO3 | 1.016 (0.962, 1.072) | 1.089 (1.015, 1.169) | 0.890 (0.803, 0.985) |
|  | NH4 | 1.058 (1.022, 1.095) | 1.065 (1.016, 1.116) | 1.041 (0.956, 1.134) |
| The third trimester cumulative average | CO | - | - | 1.010 (0.880, 1.159) |
|  | NO2 | - | - | 1.002 (0.872, 1.152) |
|  | NOx | - | - | 0.958 (0.886, 1.035) |
|  | O3 | - | - | 1.122 (1.012, 1.244) |
|  | PM10 | - | - | 1.000 (0.956, 1.045) |
|  | PM2.5 | - | - | 0.988 (0.924, 1.055) |
|  | EC | - | - | 0.996 (0.913, 1.087) |
|  | OC | - | - | 0.961 (0.850, 1.087) |
|  | NO3 | - | - | 0.971 (0.898, 1.049) |
|  | NH4 | - | - | 0.963 (0.870, 1.066) |

**Table S4**. Hazard ratio (HR) estimates per unit IQR increase and 95% CI's for all ten pollutants during five exposure windows (the first month, weekly exposure, the first trimester exposure, the second trimester exposure, and the third trimester exposure) for stillbirths that happened after the 37^th^ week of pregnancy. Estimates are adjusted for individual-level maternal age, marital status, race & ethnicity, conception year, and tract-level median household income.

| Exposure time window | Pollutants | Stillbirth after the 37^th^ week |
| --- | --- | --- |
| The first month | CO | 0.860 (0.682, 1.085) |
|  | NO2 | 0.879 (0.702, 1.100) |
|  | NOx | 0.949 (0.794, 1.134) |
|  | O3 | 0.962 (0.814, 1.138) |
|  | PM10 | 0.930 (0.811, 1.066) |
|  | PM2.5 | 0.953 (0.850, 1.068) |
|  | EC | 0.831 (0.647, 1.067) |
|  | OC | 0.942 (0.777, 1.143) |
|  | NO3 | 0.934 (0.828, 1.054) |
|  | NH4 | 0.998 (0.873, 1.141) |
| Weekly | CO | 1.068 (0.896, 1.273) |
|  | NO2 | 1.029 (0.839, 1.261) |
|  | NOx | 1.028 (0.937, 1.129) |
|  | O3 | 1.122 (0.947, 1.328) |
|  | PM10 | 1.019 (0.917, 1.133) |
|  | PM2.5 | 1.042 (0.931, 1.167) |
|  | EC | 1.032 (0.939, 1.134) |
|  | OC | 0.972 (0.863, 1.094) |
|  | NO3 | 0.977 (0.880, 1.085) |
|  | NH4 | 0.988 (0.892, 1.095) |
| The first trimester cumulative average | CO | 0.933 (0.731, 1.190) |
|  | NO2 | 0.942 (0.744, 1.192) |
|  | NOx | 1.005 (0.859, 1.175) |
|  | O3 | 1.060 (0.813, 1.383) |
|  | PM10 | 1.146 (0.874, 1.503) |
|  | PM2.5 | 1.087 (0.874, 1.353) |
|  | EC | 0.884 (0.712, 1.099) |
|  | OC | 0.938 (0.703, 1.253) |
|  | NO3 | 0.776 (0.643, 0.936) |
|  | NH4 | 1.034 (0.773, 1.383) |
| The second trimester cumulative average | CO | 0.838 (0.513, 1.369) |
|  | NO2 | 1.552 (1.193, 2.019) |
|  | NOx | 1.104 (0.894, 1.362) |
|  | O3 | 1.026 (0.739, 1.424) |
|  | PM10 | 1.062 (0.858, 1.313) |
|  | PM2.5 | 0.966 (0.799, 1.168) |
|  | EC | 1.118 (0.925, 1.350) |
|  | OC | 0.800 (0.547, 1.172) |
|  | NO3 | 0.867 (0.694, 1.082) |
|  | NH4 | 1.011 (0.840, 1.218) |
| The third trimester cumulative average | CO | 0.728 (0.373, 1.422) |
|  | NO2 | 0.767 (0.471, 1.250) |
|  | NOx | 0.756 (0.511, 1.118) |
|  | O3 | 1.349 (0.815, 2.232) |
|  | PM10 | 1.135 (0.975, 1.321) |
|  | PM2.5 | 1.055 (0.850, 1.309) |
|  | EC | 0.977 (0.668, 1.429) |
|  | OC | 0.889 (0.629, 1.256) |
|  | NO3 | 1.209 (0.946, 1.545) |
|  | NH4 | 1.127 (0.872, 1.457) |

**Table S5**. Hazard ratio (HR) estimates per unit IQR increase and 95% CI's for all ten pollutants during five exposure windows (the first month, weekly exposure, the first trimester exposure, the second trimester exposure, and the third trimester exposure) for all stillbirths, stillbirths that happened in the second trimester, and stillbirths that happened in the third trimester, adjusting for weekly average maximum temperature. Estimates are adjusted for individual-level maternal age, marital status, race & ethnicity, conception year, and tract-level median household income.

| Exposure time window | Pollutants | All stillbirth (n=8384) | 2nd trimester (n=4670) | 3rd trimester (n=3714) |
| --- | --- | --- | --- | --- |
| The first month | CO | 1.011 (0.952, 1.074) | 1.066 (0.968, 1.175) | 0.933 (0.850, 1.024) |
|  | NO2 | 0.974 (0.938, 1.010) | 1.020 (0.954, 1.091) | 0.909 (0.846, 0.977) |
|  | NOx | 0.985 (0.955, 1.016) | 0.995 (0.952, 1.040) | 0.966 (0.920, 1.015) |
|  | O3 | 0.941 (0.888, 0.997) | 0.934 (0.844, 1.033) | 0.948 (0.854, 1.052) |
|  | PM10 | 0.980 (0.952, 1.009) | 1.019 (0.966, 1.076) | 0.923 (0.865, 0.984) |
|  | PM2.5 | 0.982 (0.953, 1.011) | 1.013 (0.951, 1.078) | 0.941 (0.884, 1.000) |
|  | EC | 0.970 (0.942, 1.000) | 1.003 (0.951, 1.057) | 0.921 (0.850, 0.997) |
|  | OC | 0.997 (0.949, 1.047) | 1.020 (0.944, 1.102) | 0.970 (0.884, 1.064) |
|  | NO3 | 1.019 (0.986, 1.052) | 1.041 (0.981, 1.104) | 1.000 (0.927, 1.079) |
|  | NH4 | 1.004 (0.970, 1.038) | 1.054 (0.973, 1.142) | 0.940 (0.873, 1.011) |
| Weekly | CO | 1.076 (1.024, 1.131) | 1.152 (1.054, 1.260) | 0.978 (0.912, 1.050) |
|  | NO2 | 1.039 (0.998, 1.081) | 1.112 (1.042, 1.188) | 0.947 (0.872, 1.028) |
|  | NOx | 1.024 (0.995, 1.054) | 1.072 (1.019, 1.128) | 0.959 (0.914, 1.007) |
|  | O3 | 1.059 (0.999, 1.122) | 1.076 (0.983, 1.178) | 1.046 (0.981, 1.115) |
|  | PM10 | 1.039 (0.996, 1.084) | 1.076 (1.028, 1.127) | 0.997 (0.929, 1.069) |
|  | PM2.5 | 1.016 (0.974, 1.058) | 1.056 (1.001, 1.115) | 0.972 (0.916, 1.031) |
|  | EC | 1.028 (0.980, 1.078) | 1.069 (1.002, 1.141) | 0.983 (0.914, 1.058) |
|  | OC | 1.018 (0.964, 1.075) | 1.043 (0.980, 1.111) | 0.986 (0.917, 1.060) |
|  | NO3 | 1.016 (0.982, 1.050) | 1.050 (1.003, 1.100) | 0.979 (0.922, 1.040) |
|  | NH4 | 1.023 (0.982, 1.065) | 1.060 (1.007, 1.116) | 0.987 (0.929, 1.048) |
| The first trimester cumulative average | CO | 1.058 (0.980, 1.142) | 1.103 (0.978, 1.245) | 0.995 (0.889, 1.114) |
|  | NO2 | 1.004 (0.949, 1.063) | 1.052 (0.958, 1.157) | 0.942 (0.864, 1.028) |
|  | NOx | 1.003 (0.954, 1.055) | 1.026 (0.956, 1.100) | 0.967 (0.895, 1.046) |
|  | O3 | 0.962 (0.911, 1.016) | 0.926 (0.830, 1.033) | 1.017 (0.897, 1.153) |
|  | PM10 | 0.992 (0.953, 1.033) | 1.036 (0.952, 1.127) | 0.930 (0.838, 1.032) |
|  | PM2.5 | 0.997 (0.971, 1.025) | 1.035 (0.968, 1.105) | 0.947 (0.875, 1.024) |
|  | EC | 0.990 (0.934, 1.050) | 1.024 (0.930, 1.128) | 0.944 (0.862, 1.033) |
|  | OC | 1.033 (0.976, 1.092) | 1.032 (0.932, 1.142) | 1.037 (0.927, 1.159) |
|  | NO3 | 1.022 (0.986, 1.059) | 1.088 (1.020, 1.161) | 0.944 (0.861, 1.034) |
|  | NH4 | 1.014 (0.985, 1.045) | 1.073 (0.986, 1.168) | 0.942 (0.857, 1.035) |
| The second trimester cumulative average | CO | 1.077 (0.975, 1.189) | 1.126 (0.995, 1.274) | 0.975 (0.816, 1.166) |
|  | NO2 | 1.064 (1.001, 1.131) | 1.120 (1.033, 1.215) | 0.963 (0.845, 1.098) |
|  | NOx | 1.026 (0.983, 1.070) | 1.084 (0.998, 1.178) | 0.918 (0.794, 1.062) |
|  | O3 | 1.047 (0.969, 1.133) | 1.063 (0.973, 1.162) | 1.030 (0.918, 1.157) |
|  | PM10 | 1.074 (1.039, 1.109) | 1.092 (1.046, 1.141) | 1.030 (0.972, 1.091) |
|  | PM2.5 | 1.045 (1.006, 1.085) | 1.068 (1.011, 1.128) | 0.994 (0.932, 1.060) |
|  | EC | 1.044 (0.995, 1.096) | 1.079 (1.002, 1.161) | 0.984 (0.882, 1.097) |
|  | OC | 1.022 (0.967, 1.081) | 1.091 (1.012, 1.177) | 0.907 (0.812, 1.013) |
|  | NO3 | 1.019 (0.966, 1.075) | 1.093 (1.011, 1.182) | 0.892 (0.806, 0.988) |
|  | NH4 | 1.058 (1.022, 1.095) | 1.065 (1.016, 1.116) | 1.040 (0.956, 1.132) |
| The third trimester cumulative average | CO | - | - | 1.006 (0.875, 1.156) |
|  | NO2 | - | - | 1.003 (0.873, 1.152) |
|  | NOx | - | - | 0.958 (0.887, 1.035) |
|  | O3 | - | - | 1.119 (1.007, 1.243) |
|  | PM10 | - | - | 0.996 (0.948, 1.046) |
|  | PM2.5 | - | - | 0.984 (0.921, 1.051) |
|  | EC | - | - | 0.993 (0.907, 1.087) |
|  | OC | - | - | 0.959 (0.846, 1.087) |
|  | NO3 | - | - | 0.977 (0.896, 1.066) |
|  | NH4 | - | - | 0.963 (0.870, 1.066) |

**Table S6**. Hazard ratio (HR) estimates per unit IQR increase and 95% CI's for all ten pollutants during five exposure windows (the first month, weekly exposure, the first trimester exposure, the second trimester exposure, and the third trimester exposure) for all stillbirths, stillbirths that happened in the second trimester, and stillbirths that happened in the third trimester, adjusting for weekly average minimum temperature. Estimates are adjusted for individual-level maternal age, marital status, race & ethnicity, conception year, and tract-level median household income.

| Exposure time window | Pollutants | All stillbirth (n=8384) | 2nd trimester (n=4670) | 3rd trimester (n=3714) |
| --- | --- | --- | --- | --- |
| The first month | CO | 1.018 (0.957, 1.082) | 1.074 (0.974, 1.185) | 0.938 (0.852, 1.032) |
|  | NO2 | 0.973 (0.938, 1.009) | 1.019 (0.954, 1.088) | 0.908 (0.845, 0.977) |
|  | NOx | 0.985 (0.955, 1.017) | 0.996 (0.952, 1.041) | 0.966 (0.920, 1.015) |
|  | O3 | 0.959 (0.909, 1.012) | 0.956 (0.873, 1.048) | 0.961 (0.862, 1.072) |
|  | PM10 | 0.993 (0.966, 1.020) | 1.033 (0.976, 1.092) | 0.935 (0.879, 0.993) |
|  | PM2.5 | 0.991 (0.964, 1.018) | 1.023 (0.965, 1.085) | 0.948 (0.890, 1.010) |
|  | EC | 0.978 (0.948, 1.009) | 1.012 (0.961, 1.066) | 0.927 (0.854, 1.006) |
|  | OC | 1.006 (0.960, 1.055) | 1.031 (0.959, 1.109) | 0.977 (0.892, 1.070) |
|  | NO3 | 1.004 (0.972, 1.037) | 1.020 (0.969, 1.074) | 0.991 (0.921, 1.066) |
|  | NH4 | 1.003 (0.969, 1.037) | 1.053 (0.972, 1.140) | 0.939 (0.872, 1.010) |
| Weekly | CO | 1.078 (1.022, 1.136) | 1.159 (1.058, 1.269) | 0.974 (0.907, 1.046) |
|  | NO2 | 1.040 (0.996, 1.085) | 1.125 (1.052, 1.202) | 0.935 (0.861, 1.016) |
|  | NOx | 1.025 (0.995, 1.056) | 1.080 (1.026, 1.137) | 0.952 (0.908, 0.998) |
|  | O3 | 1.060 (1.000, 1.123) | 1.079 (0.985, 1.182) | 1.046 (0.982, 1.113) |
|  | PM10 | 1.040 (1.000, 1.081) | 1.076 (1.030, 1.124) | 0.998 (0.933, 1.066) |
|  | PM2.5 | 1.017 (0.976, 1.059) | 1.061 (1.005, 1.119) | 0.969 (0.913, 1.027) |
|  | EC | 1.029 (0.982, 1.079) | 1.076 (1.010, 1.147) | 0.979 (0.911, 1.052) |
|  | OC | 1.019 (0.965, 1.076) | 1.048 (0.984, 1.117) | 0.981 (0.915, 1.052) |
|  | NO3 | 1.013 (0.980, 1.047) | 1.059 (1.008, 1.112) | 0.965 (0.909, 1.024) |
|  | NH4 | 1.022 (0.981, 1.065) | 1.063 (1.009, 1.121) | 0.982 (0.922, 1.045) |
| The first trimester cumulative average | CO | 1.059 (0.981, 1.144) | 1.104 (0.978, 1.247) | 0.998 (0.891, 1.118) |
|  | NO2 | 0.999 (0.941, 1.060) | 1.046 (0.948, 1.155) | 0.938 (0.857, 1.027) |
|  | NOx | 0.999 (0.945, 1.057) | 1.021 (0.945, 1.104) | 0.964 (0.891, 1.044) |
|  | O3 | 0.975 (0.925, 1.029) | 0.939 (0.843, 1.045) | 1.031 (0.907, 1.171) |
|  | PM10 | 1.004 (0.966, 1.043) | 1.047 (0.965, 1.137) | 0.941 (0.850, 1.041) |
|  | PM2.5 | 1.004 (0.977, 1.031) | 1.040 (0.973, 1.111) | 0.954 (0.880, 1.034) |
|  | EC | 0.993 (0.938, 1.052) | 1.027 (0.933, 1.130) | 0.948 (0.867, 1.036) |
|  | OC | 1.039 (0.984, 1.097) | 1.037 (0.940, 1.145) | 1.043 (0.936, 1.163) |
|  | NO3 | 1.000 (0.959, 1.043) | 1.065 (0.991, 1.144) | 0.925 (0.843, 1.014) |
|  | NH4 | 1.012 (0.980, 1.045) | 1.071 (0.983, 1.167) | 0.940 (0.855, 1.033) |
| The second trimester cumulative average | CO | 1.079 (0.976, 1.193) | 1.126 (0.993, 1.276) | 0.979 (0.819, 1.169) |
|  | NO2 | 1.068 (1.005, 1.135) | 1.128 (1.040, 1.224) | 0.963 (0.842, 1.102) |
|  | NOx | 1.028 (0.985, 1.072) | 1.089 (1.003, 1.182) | 0.917 (0.794, 1.060) |
|  | O3 | 1.048 (0.972, 1.130) | 1.061 (0.975, 1.156) | 1.035 (0.922, 1.162) |
|  | PM10 | 1.073 (1.041, 1.106) | 1.088 (1.043, 1.135) | 1.033 (0.977, 1.092) |
|  | PM2.5 | 1.045 (1.006, 1.085) | 1.066 (1.011, 1.125) | 0.997 (0.934, 1.065) |
|  | EC | 1.045 (0.997, 1.097) | 1.078 (1.002, 1.160) | 0.987 (0.890, 1.094) |
|  | OC | 1.024 (0.967, 1.084) | 1.091 (1.012, 1.176) | 0.909 (0.813, 1.017) |
|  | NO3 | 1.019 (0.967, 1.074) | 1.100 (1.014, 1.193) | 0.888 (0.800, 0.985) |
|  | NH4 | 1.059 (1.022, 1.096) | 1.066 (1.017, 1.117) | 1.041 (0.956, 1.134) |
| The third trimester cumulative average | CO | - | - | 1.010 (0.880, 1.160) |
|  | NO2 | - | - | 0.993 (0.863, 1.142) |
|  | NOx | - | - | 0.953 (0.881, 1.031) |
|  | O3 | - | - | 1.123 (1.013, 1.245) |
|  | PM10 | - | - | 1.002 (0.958, 1.049) |
|  | PM2.5 | - | - | 0.989 (0.925, 1.056) |
|  | EC | - | - | 0.995 (0.912, 1.087) |
|  | OC | - | - | 0.960 (0.850, 1.085) |
|  | NO3 | - | - | 0.962 (0.887, 1.043) |
|  | NH4 | - | - | 0.962 (0.868, 1.065) |

**Table S7.** Hazard ratios (HRs) and 95% CIs from single-pollutant and two-pollutant Cox proportional hazards models. Two pollutant models included NO_2_ adjusted for PM_2.5_, O_3_ adjusted for PM_2.5_, and PM_2.5_ adjusted for O_3_.

| Exposure time window | Pollutants | All stillbirth (n=8384) | 2nd trimester (4670) | 3rd trimester (3714) |
| --- | --- | --- | --- | --- |
| The first month | NO2 | 0.973 (0.938, 1.009) | 1.019 (0.954, 1.088) | 0.908 (0.845, 0.977) |
|  | NO2 + PM2.5 | 0.975 (0.934, 1.017) | 1.009 (0.942, 1.081) | 0.926 (0.863, 0.993) |
|  | O3 | 0.959 (0.909, 1.012) | 0.956 (0.873, 1.048) | 0.961 (0.862, 1.072) |
|  | O3 + PM2.5 | 0.953 (0.888, 1.022) | 0.915 (0.822, 1.019) | 1.004 (0.884, 1.141) |
|  | PM2.5 | 0.991 (0.964, 1.018) | 1.023 (0.965, 1.085) | 0.948 (0.890, 1.010) |
|  | PM2.5 + O3 | 1.009 (0.971, 1.047) | 1.057 (0.987, 1.132) | 0.947 (0.880, 1.018) |
| Weekly | NO2 | 1.038 (0.997, 1.081) | 1.112 (1.041, 1.188) | 0.946 (0.870, 1.029) |
|  | NO2 + PM2.5 | 1.033 (0.992, 1.075) | 1.089 (1.013, 1.170) | 0.956 (0.887, 1.030) |
|  | O3 | 1.059 (1.001, 1.122) | 1.075 (0.983, 1.176) | 1.049 (0.987, 1.114) |
|  | O3 + PM2.5 | 1.069 (1.005, 1.137) | 1.044 (0.947, 1.151) | 1.103 (1.017, 1.197) |
|  | PM2.5 | 1.016 (0.976, 1.058) | 1.056 (1.001, 1.114) | 0.973 (0.919, 1.031) |
|  | PM2.5 + O3 | 0.989 (0.947, 1.033) | 1.038 (0.983, 1.096) | 0.935 (0.868, 1.007) |
| The first trimester cumulative average | NO2 | 1.002 (0.947, 1.059) | 1.050 (0.958, 1.151) | 0.939 (0.861, 1.025) |
|  | NO2 + PM2.5 | 1.001 (0.944, 1.061) | 1.040 (0.948, 1.139) | 0.952 (0.876, 1.035) |
|  | O3 | 0.974 (0.924, 1.026) | 0.937 (0.842, 1.042) | 1.030 (0.905, 1.173) |
|  | O3 + PM2.5 | 0.964 (0.897, 1.035) | 0.888 (0.792, 0.996) | 1.086 (0.932, 1.266) |
|  | PM2.5 | 1.002 (0.975, 1.030) | 1.037 (0.970, 1.110) | 0.954 (0.879, 1.036) |
|  | PM2.5 + O3 | 1.015 (0.974, 1.057) | 1.079 (1.004, 1.159) | 0.928 (0.841, 1.024) |
| The second trimester cumulative average | NO2 | 1.064 (1.002, 1.131) | 1.121 (1.033, 1.215) | 0.963 (0.845, 1.097) |
|  | NO2 + PM2.5 | 1.038 (0.975, 1.105) | 1.081 (0.978, 1.195) | 0.958 (0.830, 1.106) |
|  | O3 | 1.049 (0.972, 1.131) | 1.062 (0.975, 1.156) | 1.035 (0.921, 1.163) |
|  | O3 + PM2.5 | 1.020 (0.936, 1.113) | 1.017 (0.922, 1.122) | 1.040 (0.909, 1.190) |
|  | PM2.5 | 1.045 (1.006, 1.085) | 1.067 (1.011, 1.125) | 0.997 (0.933, 1.066) |
|  | PM2.5 + O3 | 1.035 (0.993, 1.078) | 1.054 (0.993, 1.119) | 0.987 (0.912, 1.069) |
| The third trimester cumulative average | NO2 | - | - | 1.002 (0.872, 1.152) |
|  | NO2 + PM2.5 | - | - | 1.005 (0.859, 1.177) |
|  | O3 | - | - | 1.122 (1.012, 1.244) |
|  | O3 + PM2.5 | - | - | 1.174 (1.012, 1.362) |
|  | PM2.5 | - | - | 0.988 (0.924, 1.055) |
|  | PM2.5 + O3 | - | - | 0.932 (0.848, 1.025) |

**Table S8**. Hazard ratio (HR) estimates per unit IQR increase and 95% CIs for all ten pollutants for stillbirths occurring between the 37^th^ to the 42^nd^ gestational week (full term) and stillbirths occurring before the 37^th^ gestational week.

| Exposure time window | Pollutants | 37 to 42 weeks | Less than 37 weeks |
| --- | --- | --- | --- |
| The first month | CO | 0.851 (0.709, 1.021) | 1.030 (0.948, 1.118) |
|  | NO2 | 0.825 (0.719, 0.945) | 0.999 (0.919, 1.086) |
|  | NOx | 0.932 (0.849, 1.024) | 1.000 (0.939, 1.065) |
|  | O3 | 0.959 (0.841, 1.093) | 1.004 (0.914, 1.104) |
|  | PM10 | 0.927 (0.847, 1.014) | 1.028 (0.974, 1.084) |
|  | PM2.5 | 0.936 (0.852, 1.029) | 0.992 (0.933, 1.055) |
|  | EC | 0.842 (0.709, 1.000) | 1.016 (0.955, 1.080) |
|  | OC | 0.911 (0.762, 1.090) | 0.996 (0.932, 1.064) |
|  | NO3 | 0.850 (0.752, 0.961) | 1.049 (0.983, 1.118) |
|  | NH4 | 0.962 (0.859, 1.078) | 1.055 (0.982, 1.134) |
| Weekly | CO | 1.014 (0.855, 1.203) | 1.051 (0.990, 1.115) |
|  | NO2 | 1.028 (0.871, 1.213) | 1.004 (0.914, 1.103) |
|  | NOx | 1.038 (0.966, 1.116) | 1.013 (0.953, 1.078) |
|  | O3 | 1.092 (0.990, 1.203) | 1.057 (0.971, 1.150) |
|  | PM10 | 0.995 (0.907, 1.091) | 1.036 (0.995, 1.079) |
|  | PM2.5 | 1.012 (0.925, 1.108) | 1.013 (0.960, 1.069) |
|  | EC | 1.032 (0.936, 1.137) | 1.021 (0.953, 1.093) |
|  | OC | 0.946 (0.854, 1.048) | 1.038 (0.980, 1.099) |
|  | NO3 | 1.039 (0.966, 1.117) | 1.025 (0.976, 1.077) |
|  | NH4 | 0.977 (0.873, 1.093) | 1.011 (0.961, 1.063) |
| The first trimester cumulative average | CO | 0.878 (0.708, 1.089) | 1.024 (0.907, 1.157) |
|  | NO2 | 0.868 (0.740, 1.018) | 1.036 (0.969, 1.107) |
|  | NOx | 0.941 (0.834, 1.061) | 1.010 (0.946, 1.078) |
|  | O3 | 1.042 (0.885, 1.228) | 1.003 (0.877, 1.147) |
|  | PM10 | 1.056 (0.847, 1.317) | 1.088 (1.023, 1.157) |
|  | PM2.5 | 1.036 (0.867, 1.238) | 1.029 (0.967, 1.095) |
|  | EC | 0.885 (0.753, 1.041) | 1.028 (0.961, 1.100) |
|  | OC | 0.906 (0.699, 1.175) | 1.065 (0.969, 1.171) |
|  | NO3 | 0.740 (0.630, 0.868) | 1.091 (0.984, 1.211) |
|  | NH4 | 0.993 (0.801, 1.231) | 1.078 (1.002, 1.160) |
| The second trimester cumulative average | CO | 0.830 (0.584, 1.180) | 1.088 (0.948, 1.248) |
|  | NO2 | 1.276 (0.971, 1.676) | 1.001 (0.911, 1.100) |
|  | NOx | 1.094 (0.893, 1.341) | 1.013 (0.953, 1.077) |
|  | O3 | 1.042 (0.834, 1.303) | 1.088 (0.999, 1.184) |
|  | PM10 | 1.098 (0.933, 1.292) | 1.076 (1.016, 1.140) |
|  | PM2.5 | 1.001 (0.874, 1.146) | 1.070 (1.001, 1.143) |
|  | EC | 1.017 (0.846, 1.222) | 1.046 (0.939, 1.164) |
|  | OC | 0.770 (0.575, 1.032) | 1.082 (1.008, 1.162) |
|  | NO3 | 0.914 (0.758, 1.104) | 1.050 (1.010, 1.092) |
|  | NH4 | 1.011 (0.855, 1.196) | 1.066 (0.990, 1.149) |
| The third trimester cumulative average | CO | 0.899 (0.560, 1.444) | 1.031 (0.828, 1.284) |
|  | NO2 | 0.855 (0.601, 1.217) | 1.030 (0.833, 1.274) |
|  | NOx | 0.928 (0.716, 1.203) | 1.047 (0.877, 1.250) |
|  | O3 | 1.127 (0.799, 1.589) | 1.099 (0.926, 1.306) |
|  | PM10 | 1.057 (0.893, 1.251) | 0.926 (0.851, 1.007) |
|  | PM2.5 | 1.028 (0.810, 1.305) | 0.904 (0.816, 1.002) |
|  | EC | 1.012 (0.712, 1.436) | 0.956 (0.850, 1.077) |
|  | OC | 0.819 (0.658, 1.019) | 1.007 (0.847, 1.198) |
|  | NO3 | 1.223 (1.029, 1.453) | 0.904 (0.802, 1.018) |
|  | NH4 | 1.146 (0.911, 1.442) | 0.787 (0.686, 0.903) |

**Figure S1**. Hazard ratio (HR) estimates per unit IQR increase and 95% CIs for all ten pollutants during all five exposure windows restricted to stillbirths occurring after the 37^th^ week of pregnancy.

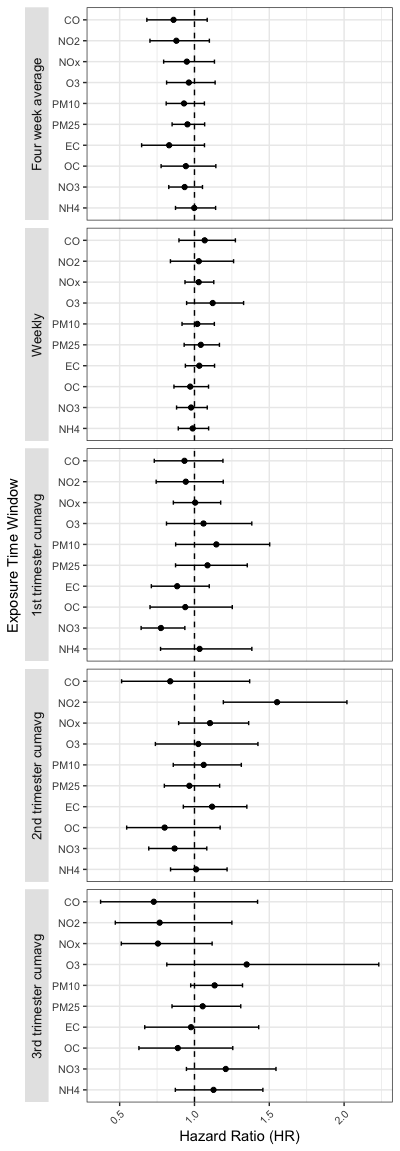

**Figure S2**. Comparison of hazard ratio (HR) estimates per unit IQR increase and 95% CIs for all ten pollutants during all exposure windows (the first week, weekly the first trimester, the second trimester, and the third trimester exposure) for all stillbirths, stillbirths that happened in the second trimester, and stillbirths that happened in the third trimester between main analysis and sensitivity analysis adjusting for weekly average maximum temperature.

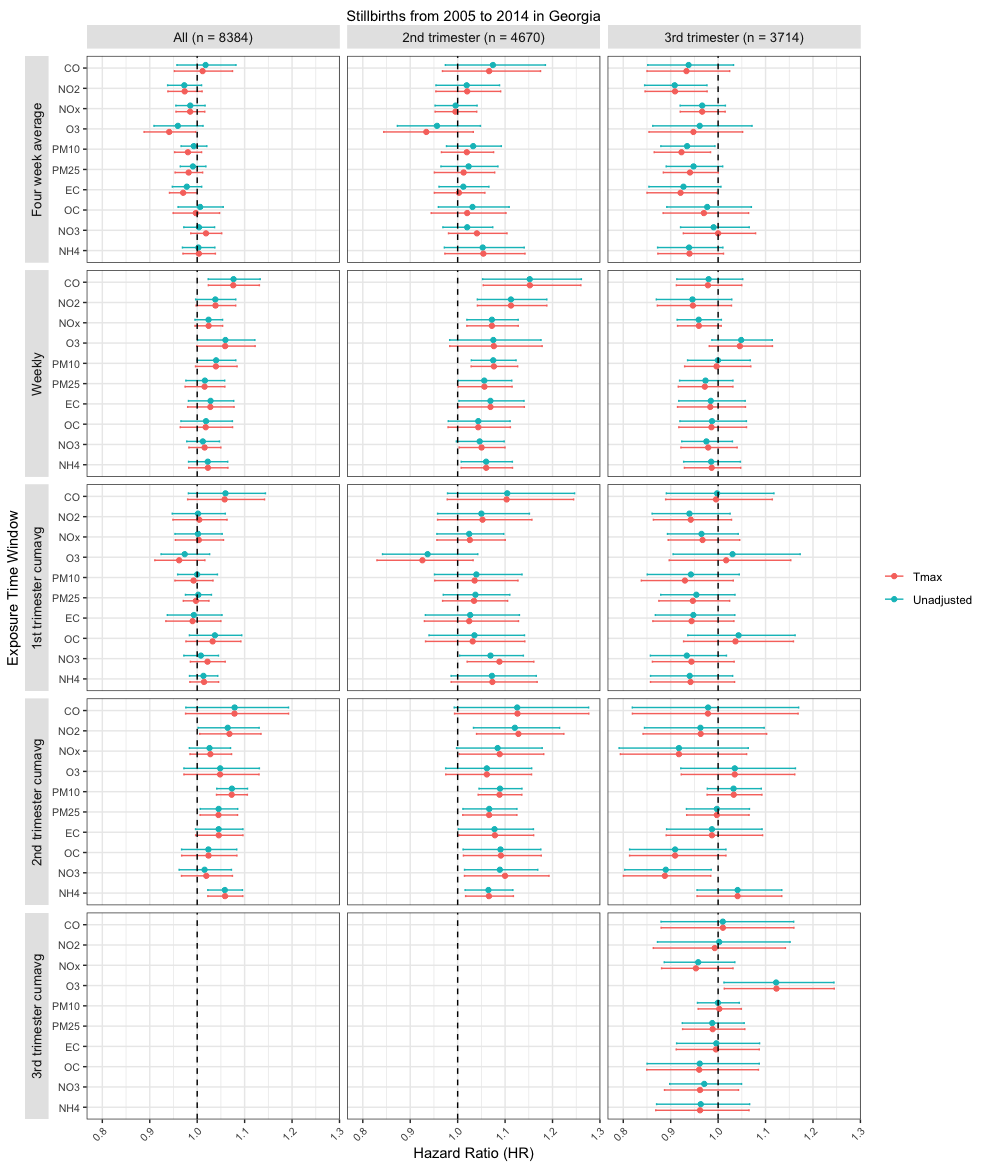

**Figure S3**. Comparison of hazard ratio (HR) estimates per unit IQR increase and 95% CI's for all ten pollutants during all exposure windows (the first week, weekly the first trimester, the second trimester, and the third trimester exposure) for all stillbirths, stillbirths that happened in the second trimester, and stillbirths that happened in the third trimester between main analysis and sensitivity analysis adjusting for weekly average minimum temperature.

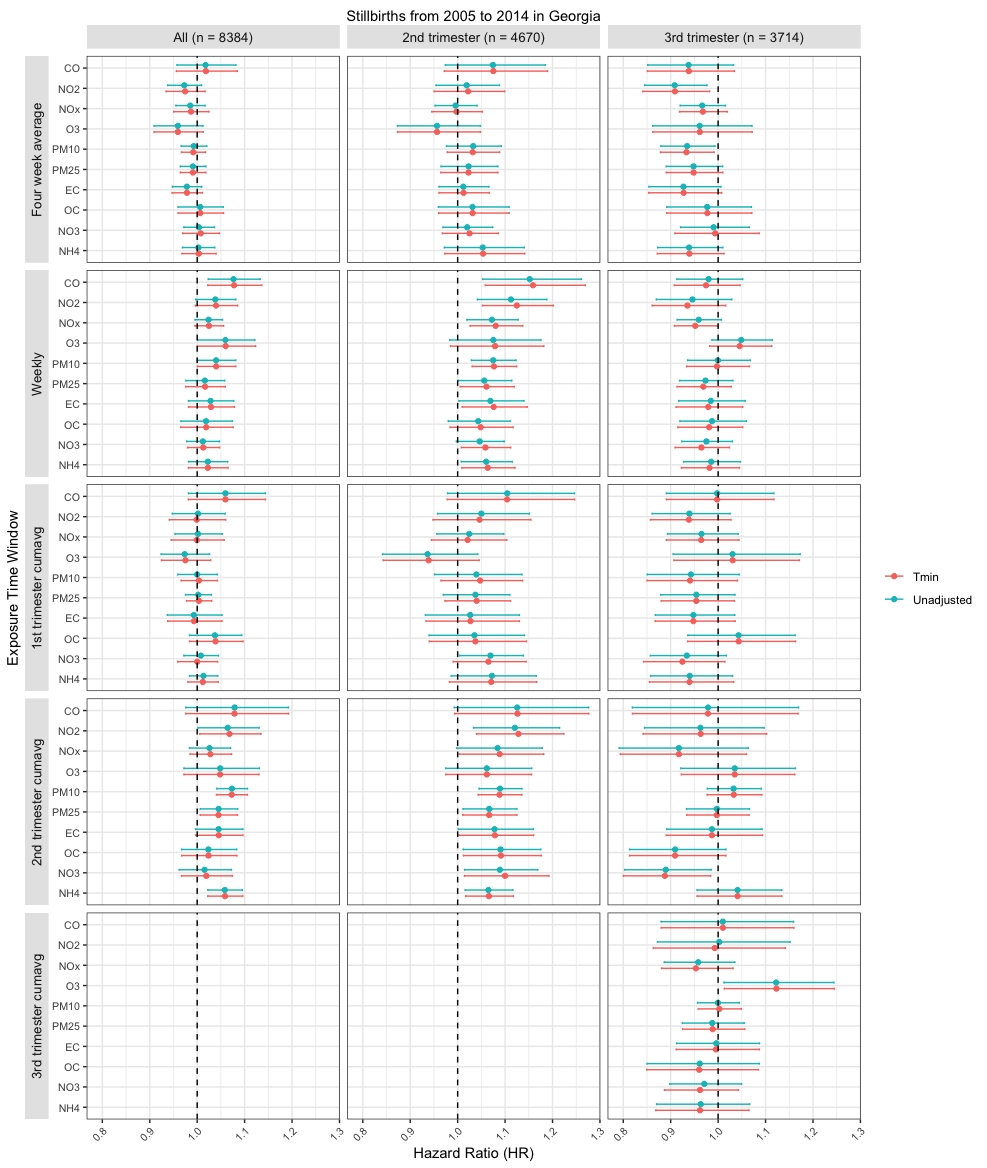

**Figure S4**. Comparison of hazard ratio (HR) estimates per unit IQR increase and 95% CI's for single-pollutant and two-pollutant model estimates for NO_2_, O_3_, and PM_2.5_. Hazard ratios (HRs) and 95% confidence intervals correspond to an interquartile range increase in pollutant exposure. Two-pollutant models adjusted NO_2_ for PM_2.5_, O_3_ for PM_2.5_, and PM_2.5_ for O_3_.

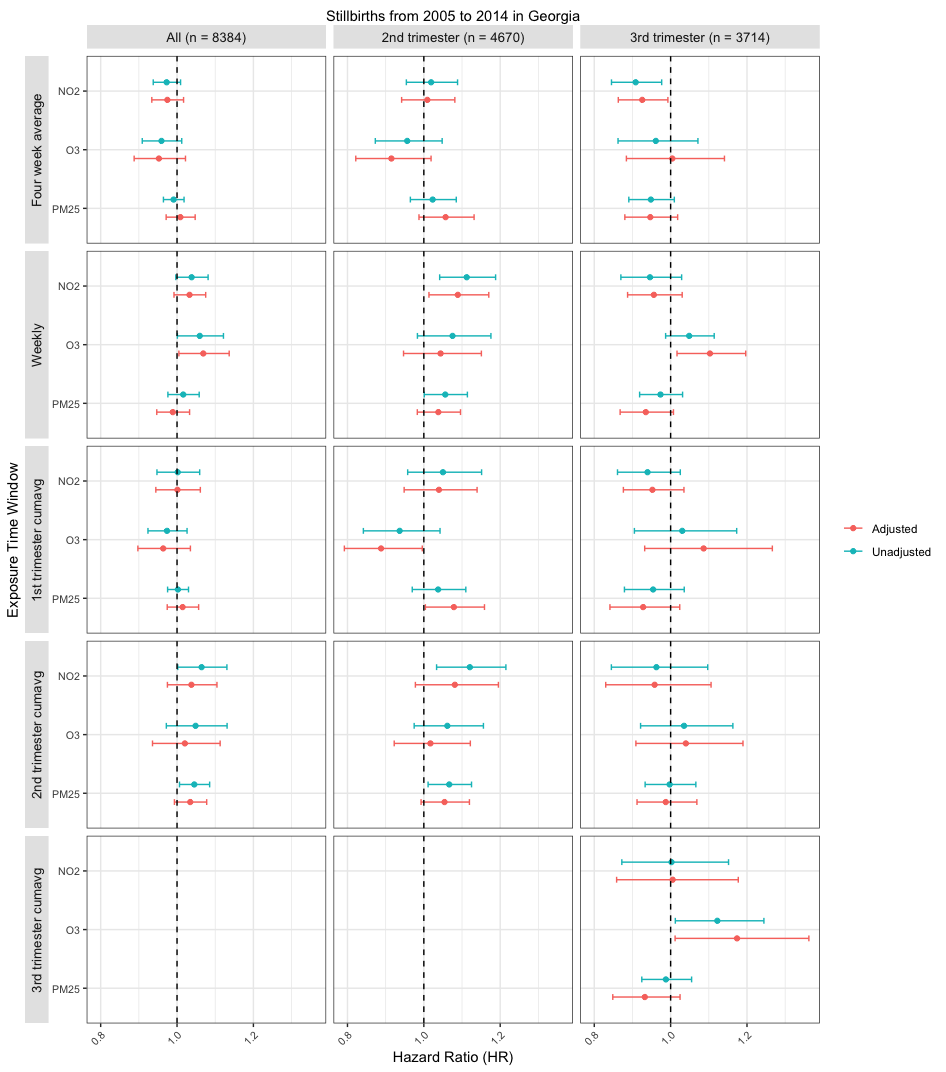

**Figure S5**. Comparison of hazard ratio (HR) estimates per unit IQR increase and 95% CIs for all ten pollutants during all exposure windows between stillbirths that happened from the 37^th^ to the 42^nd^ gestational week (full term) and stillbirths occurring before the 37^th^ gestational week.

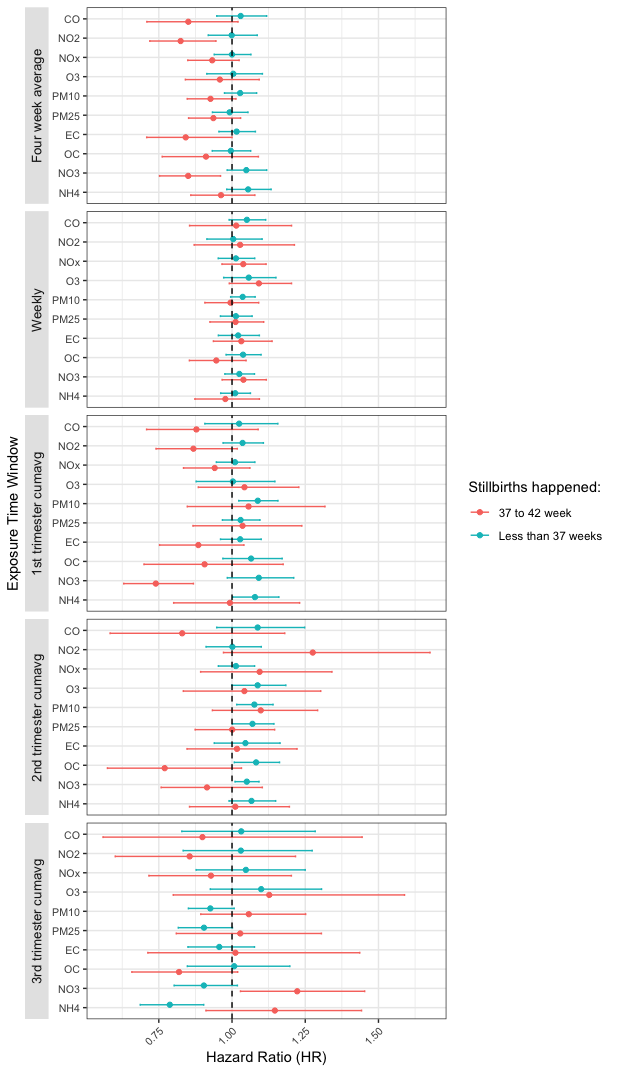
